# An analysis of retinal safety when using a Laser based low-level red light therapy device for myopia

**DOI:** 10.64898/2026.05.05.26352503

**Authors:** Karl Schulmeister, John Marshall

**Author notes:** **Corresponding Author** Karl Schulmeister, Ph.D., Seibersdorf Labor GmbH, 2444 Seibersdorf, Austria.

## Abstract

**Purpose:** To evaluate the retinal safety of repeated low-level red-light (RLRL) therapy using the Eyerising Myopia Management Device (EMMD) by analysing exposure parameters relative to established thermal and photochemical retinal injury thresholds and empirical human exposure data.

**Methods:** Emission characteristics of the EMMD were measured in an accredited laboratory under worst-case conditions. Parameters assessed included wavelength, intraocular power, corneal irradiance, and retinal image characteristics across accommodative states. These measurements were compared with international safety standards, maximum permissible exposure limits, and experimentally derived retinal injury thresholds from animal studies and validated computational models. The effects of repeated exposures from RLRL therapy using the EMMD were evaluated using photochemical additivity principles and repair kinetics, and further contextualised using human volunteer exposure data.

**Results:** The EMMD emitted red laser radiation at 654–655 nm with a maximum intraocular power of approximately 1 mW through a 7 mm pupil, placing it within Class 3R and marginally above the Class 2 limit. Corneal irradiance was approximately 26 W m^− 2^, well below conservative photochemical exposure limits. Thermal injury modelling indicated retinal damage thresholds above device exposure, including under worst-case assumptions of minimal retinal image size and absence of eye movements. Accounting for repeated daily exposures and photochemical additivity, safety margins remained approximately 3-fold for a 7 mm pupil and approximately 8-fold for a more realistic 4 mm pupil. Human volunteer studies demonstrated no detectable structural or functional retinal injury at exposure levels approximately five times higher than those produced by the EMMD.

**Conclusion:** Exposure parameters of RLRL therapy using the EMMD remain well below conservative retinal injury thresholds under prescribed use conditions. Integration of experimental, modelling, and human data indicates substantial safety margins, supporting its safe clinical use.

## 1 INTRODUCTION

Myopia is a common refractive condition worldwide with some evidence suggesting its prevalence may be increasing due to behavioural changes in younger populations, particularly increased near work and screen use.(1, 2) Its progression is most pronounced in childhood where increases in severity and progress may lead to high myopia. It is associated with a higher risk of various ocular complications in later life. The prevalence of myopia and increasing numbers especially in Asia are a significant public health challenge.(3)

Over the past several years, a number of treatment strategies and devices have emerged to allow repeated exposures to low-level red-light (RLRL) with the aim of slowing myopia progression in children.(4). The Eyerising Myopia Management Device (EMMD) is one such device. It was the first device to have been adopted outside of China, where this technology originated. While some systems use red light-emitting diodes, the Eyerising device employs two laser beams emitting red light at wavelengths of approximately 650 ±10 nm. Children are instructed to look into the device for three minutes, twice daily, five days per week. While several clinical studies have reported reduction in myopia progression with RLRL therapy, including systematic reviews and multicentre studies, (4-7) questions regarding retinal safety have also been raised in the literature. (8, 9)

The first priority of medicine is to ensure that interventions do no harm; therefore, rigorous safety assessment of devices used for myopia treatment is essential. Several publications have reported measurements of optical power emitted by such devices and have discussed their potential to induce retinal injury.(8-11) Over the past five decades, an extensive body of data has been generated assessing the potential for laser radiation to induce retinal injury. These studies, conducted primarily in animal models (12), and in enucleated human eyes (13, 14), examined the dependence of retinal injury threshold on wavelengths, pulse durations, retinal spot sizes and energy levels. From these experiments, injury thresholds corresponding to the energy levels required to produce irreversible retinal damage in 50% of exposures (ED50) were determined (15). Safety standards subsequently incorporated substantial safety margins to establish maximum permissible exposure (MPE) limits in the codes of practice. There is therefore a significant margin between the MPE and the measured threshold for damage.

While the empirical data base remains constant, different regulatory standards such as those developed by the American National Standard Institute (ANSI) and that of the International Electrotechnical Commission (IEC) incorporate slightly different safety margins.(16, 17) In reality, these codes of practice are highly technical documents that can be difficult for non-experts to interpret. They define criteria for retinal injury according to exposure parameters such as wavelength, retinal spot sizes, pulse durations and the resulting retinal irradiance. The MPE limits incorporate these factors and reflect the different damage mechanisms associated with varying exposure conditions.

The mechanism of retinal injury depends largely on the duration and wavelength of exposure(18-20). Extremely short pulses (nanoseconds or less) may produce thermoacoustic damage due to rapid energy disposition within the tissue. Exposures in the millisecond range primarily result in thermal injury caused by absorption of optical energy by retinal structures. Of particular relevance are exposures to short wavelength visible light, especially in the blue spectral region, where photons can induce photochemical changes in retinal tissue. This mechanism underlies the well-known blue-light hazard (peaking at approximately 440 nm) whereby exposures lasting from milliseconds to minutes, or repeated exposures over time, may selectively damage photoreceptor cells and lead to visual defects. However, this mechanism is not a significant consideration for red light devices currently used in myopia treatment.

There has been increasing clinical and public concern regarding the safety of repeated low-level red-light therapy. Particular concern has been expressed relating to systems using low-level lasers. This has to great extent arisen because of a misunderstanding with the public perception of lasers as potentially hazardous systems. At the level used in these treatment devices, it should be understood that from a biophysical perspective, the interaction of optical radiation with retinal tissue is primarily determined by exposure parameters such as wavelength, irradiance, exposure duration, and retinal image characteristics, rather than the coherence of the laser radiation (16-18). Therefore, biophysically there are no intrinsic differences between the two optical sources used in such devices and when these parameters are comparable laser and LED sources are not expected to differ in their effects on biological tissue (18-20). This paper features a detailed safety analysis of a laser-based device in order to address current clinical and public concerns.

To simplify hazard evaluation for commercial laser products, classification systems have been developed that relate to MPE limits. Class 1 systems are considered intrinsically safe because of low output levels or complete containment of the laser beam. Class 2 systems have an upper emission limit of 1mW and present extremely low risk of retinal injury. Class 3R devices have an upper limit of 5 mW and also represent a very low risk of irreversible retinal damage. Class 3B systems (5 to 500 mW) are associated with moderate to high risk, whereas Class 4 systems (> 500 mW) may pose a significant hazard for irreversible retinal or skin injury.

It should be noted that these safety standards were developed primarily to protect the general public and occupational users from inadvertent exposure to laser irradiation. They were not designed in any way to apply to therapeutic uses. It must be emphasised, therefore, that while it is useful to get some indication as to the potential unexpected consequences induced by therapeutic devices, decisions as to therapeutic regimes must be determined by risk versus benefit analysis. The present paper is a review of the exposure parameters of a laser-based device, the EMMD, and the relationship between the emitted radiation and current safety limits. For further clarification, the exposure parameters are discussed in relation to a published volunteer study, which reviewed post exposure ocular examination and patient subjective responses.

## 2 METHODS

### 2.1 The EMMD device and exposure regime

The main components of the EMMD are two laser modules contained within a cover, a binocularly eyepiece with an interpupillary distance adjustment control, a touch screen at the front of the device and a circuit housing (Fig 1).

**Figure 1.**
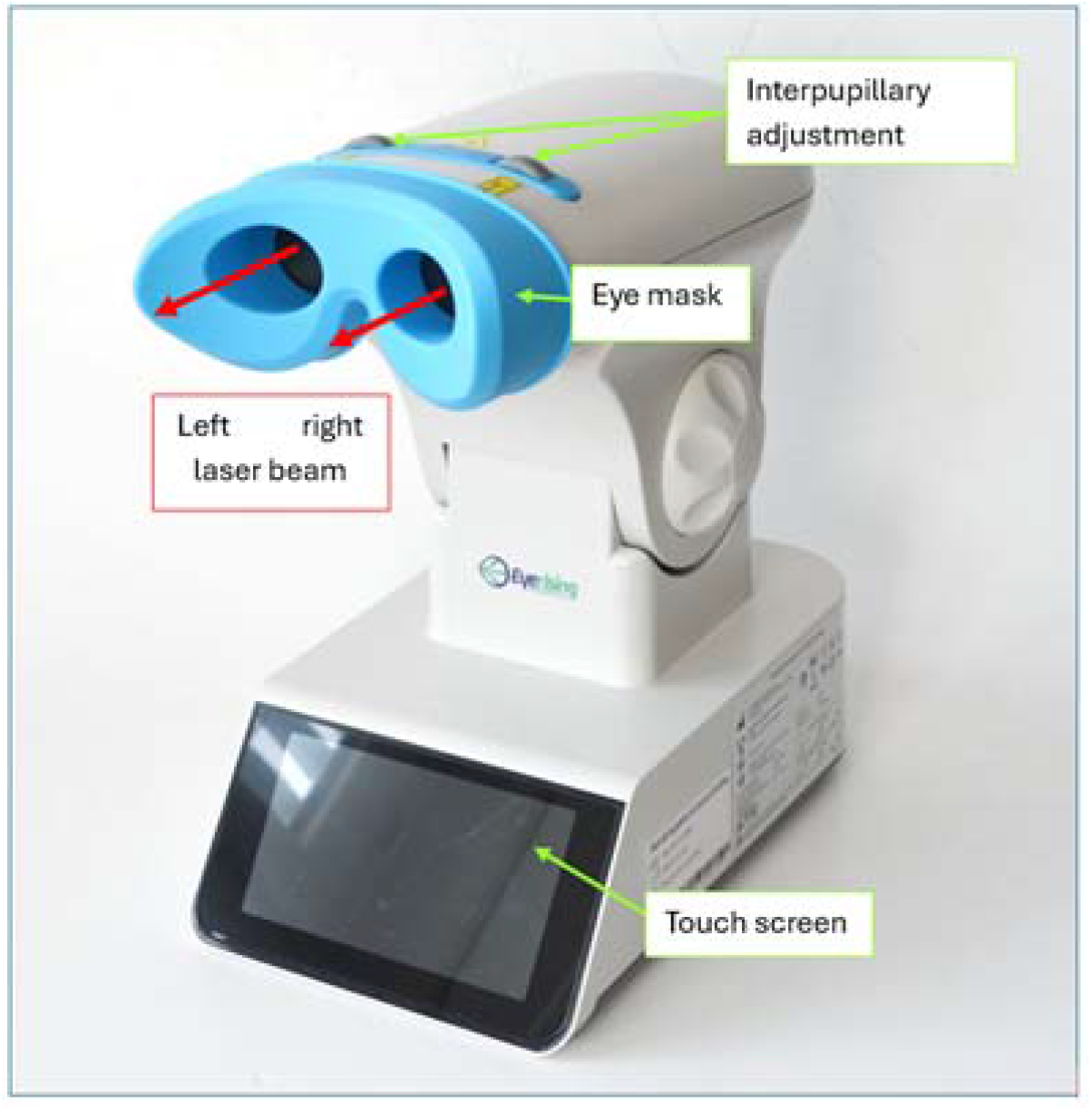
General view and main components of the EMMD.

The EMMD is operated via a touch screen. The laser emission is at a fixed level and cannot be adjusted by the user. When the laser emission is triggered via the user interface, laser radiation is emitted for a duration of 3 minutes (180 seconds), after which the emission terminates automatically. The emission cannot be restarted for 4 hours. After this interval, the emission may be restarted once more on the same day, providing a second 3-minute exposure. No further emission is possible that day. In addition, the device cannot be triggered every day, as its software only allows a maximum of 5 days in a seven-day week.

### 2.2 Measurement of the emission

Two EMMD units were examined, and the relevant exposure parameters were measured in the accredited test laboratory of Seibersdorf Labor GmbH, Austria.

The wavelength of the laser radiation was determined with a spectrometer (Flame S-XR1-ES, Ocean Optics, USA). The laser power was determined with a transient recorder (TR 9600, Gigahertz-Optik, Germany) with an integrating sphere as the input optics.

The irradiance pattern equivalent to the retinal image was determined with a laser beam profiling CCD camera (SP928, MKS Instruments, Ophir, USA). This was achieved by placing a lens with 35 mm focal length, fitted with a 7 mm aperture stop at a certain distance in front of the camera chip. By adjusting the distance between the lens and the CCD chip (the image distance), the accommodation state of this “artificial eye” could be adjusted: placing the CCD chip in the focal plane of the lens resulted in accommodation to infinity, and placing the CCD chip further away resulted in correspondingly closer accommodation distances.

The accommodation distance was varied and for each position, a CCD image was recorded. By dividing the image extent measured in µm (i.e., in the plane of the CCD chip) by the image distance in mm, the image extent was characterised in terms of angular subtense in mrad (milliradian). This approach allows the measurement to be representative of the human eye despite differences in size and focal length.

## 3 RESULTS

### 3.1 Exposure parameters

The wavelength of the lasers in the EMMD units were determined to be between 654 nm and 655 nm. In measuring the output of the device in relation to a retinal exposure an aperture of 7 mm diameter was employed, as this represents the worst-case size of the human pupil and is the standard aperture used in safety calculations. The power measured through a 7 mm aperture at the position of the patient’s eye was determined to be between 0.93 mW and 1.03 mW. For the safety assessment, a value of 1 mW was used for simplicity. The diameter of the circular laser beam at the position of the eye is larger than 7 mm and the maximum total power in the beam was measured to be 1.45 mW. Given that in practice the brightness of the beam would cause the pupil to constrict, further measurements were performed using a 4 mm aperture, which resulted in an intraocular power of 0.36 mW.

Retinal irradiance is the key determinant of risk for irreversible retinal damage and it depends on the accommodative state of the eye. The methodology used in the current study allowed characterisation of the image that is equivalent to the retinal image, with the ability to simulate accommodation in an artificial eye. Figure 2 shows the CCD image for accommodation to infinity, showing a circular profile with almost constant irradiance and sharp edges. The white square subtends 18.4 mrad (1.05 degrees), corresponding to a retinal image diameter of 313 µm.

**Figure 2.**
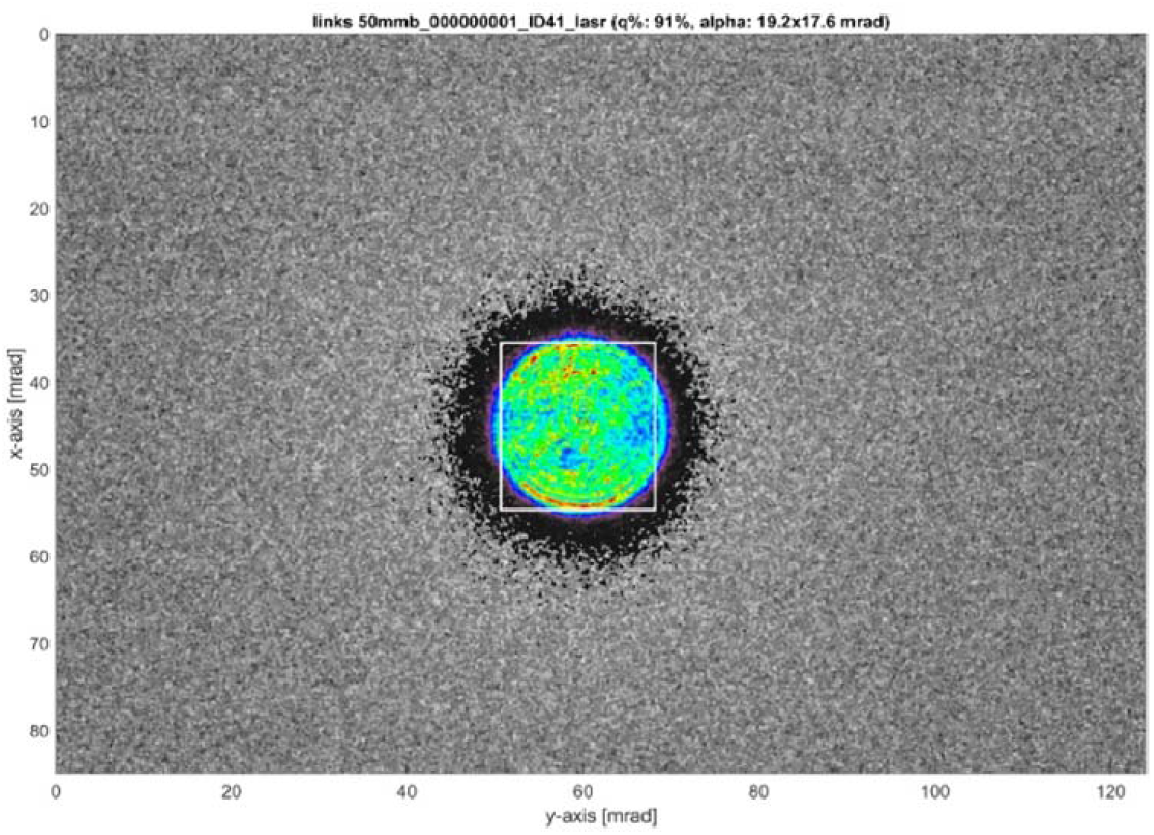
CCD image obtained for accommodation to infinity. The white triangle is the solution of an image analysis to maximise the ratio of power within the image’s section and the safety limit.

The smallest retinal image was determined for accommodation to a distance of approximately 0.32 m in front of the eye. The resulting image subtends an angle less than 1.5 mrad. In laser safety standards a value of a_min_ = 1.5 mrad is a category of laser sources referred to as ‘small sources’. This accommodation position therefore represents the worst-case condition and was used in the following analysis. This condition applies to both adults and paediatric eyes .

### 3.2 Product safety standards

For repeated low-level red-light (RLRL) therapy, currently there are no dedicated product safety standards available.

With the exception of ophthalmic instruments used for diagnosis, all laser products are subject to the international safety standard IEC 60825-1:2014. Identical national versions of this standard are enforced world-wide. IEC 60825-1 defines a classification system, ranging from Class 1 to Class 4, and related hardware safety requirements, including labelling and user information. The limits, referred to as accessible emission limits (AEL) for small sources in the red wavelength range, with the emission determined through a 7 mm aperture stop, are as follows. For Class 1: 0.39 mW, for Class 2: 1 mW, for Class 3R: 5 mW, and for Class 3B: 500 mW.

With the measured maximum emission of the EMMD of 1.03 mW, the device falls into Class 3R, although it only marginally exceeds the Class 2 limit. It should again be emphasised that IEC 60825-1 has no specific regulations for medical laser products.

Current standards, regulating the safety of optical radiation emitted by ophthalmic instruments have also not yet considered RLRL therapy. Both the USA product safety standard for ophthalmic instruments ANSI Z80.36-2021 (17) and at an international level, the equivalent standard ISO 15004-2:2024 (21) apply only to ophthalmic devices used for applications such as illumination or diagnosis and do not cover therapeutic procedures. The EMMD, being a treatment device, therefore falls outside the scope of these two standards.

## 4 DISCUSSION

### 4.1 Laser class limits

The emission limit of Class 2, namely 1 mW, is given in IEC 60825-1 to prevent thermally induced retinal injury for a minimum retinal image (small source). This is based on an assumed exposure duration of no longer than 0.25 s, representing the time that would naturally limit exposure through blinking or aversion of gaze. Cleary this assumption does not apply to an EMMD exposure as the patient is instructed to maintain fixation directly into the beam.

However, the exposure limit defined for photochemical hazard should also be considered. The maximum permissible exposure (MPE) given as corneal irradiance in Annex A of IEC 60825-1, is equal to C_3_ W m^-2^ for exposure durations longer than 100 seconds, and is defined for exposure durations up to 30 000 seconds (approximately 8 hours). This limit, expressed as constant irradiance value, does not follow the reciprocity law, under which the irradiance limit would decrease with increasing exposure duration. Instead, the limit is defined as constant irradiance because increased eye movements during longer exposures are assumed to reduce the effective retinal irradiance per unit area.

More importantly, there is a correction factor C_3_ that reflects the exponential reduction in photochemical hazard with increasing wavelength. The factor equals unity for wavelengths in the blue spectral regime, up to 450 nm, giving an MPE of 1 W m^-2^. For a wavelength of 600 nm, C_3_ = 1000 resulting in an MPE of 1 000 W m^-2^. The photochemical hazard limit is defined only up to 600 nm, reflecting the negligible photochemical hazard for wavelengths in the red spectral range, such as for the EMMD with a wavelength of 655 nm. However, as a worst-case approach, the MPE applicable for 600 nm is applied here.

The corneal irradiance associated with the EMMD is obtained by dividing 1 mW by the area of the 7 mm aperture stop, resulting in 26 W m^-2^. This value is a factor of 38 below the MPE. Thermal and photochemical mechanisms are discussed in more detail in the following sections.

### 4.2 Pupil diameter

The main numerical analysis in this paper is based on a pupil diameter of 7 mm, resulting in an intraocular power of approximately 1 mW. A 7 mm pupil is considered the maximum pupil dilation likely to be found in a young child and has therefore been adopted as a standard worst-case aperture for safety calculations. However, the luminance, or brightness, associated with viewing the laser beam is considerable. For a normally reacting eye, this will result in a pupil diameter smaller than 7 mm, which in turn reduces the intraocular power entering the eye. For accommodation to the beam waist, representing the worst-case accommodation resulting in the smallest retinal image, the retinal irradiance level will be reduced with the square of the ratio of the pupil diameters. This effect further strengthens the conclusion regarding the safety margin of the EMMD.

### 4.3 Thermal hazard threshold data

The retinal thermal hazard has been characterised using a validated computer model(22). Figure 3 shows non-human primate (NHP) threshold data (23-27) for collimated laser beams resulting in a minimum retinal spot size, for a wavelength of 633 nm as well as emissions in the green wavelength range. These empirical thresholds could be well fitted with the computer model, shown as solid lines, when assuming a retinal spot diameter of 80 µm.

**Figure 3.**
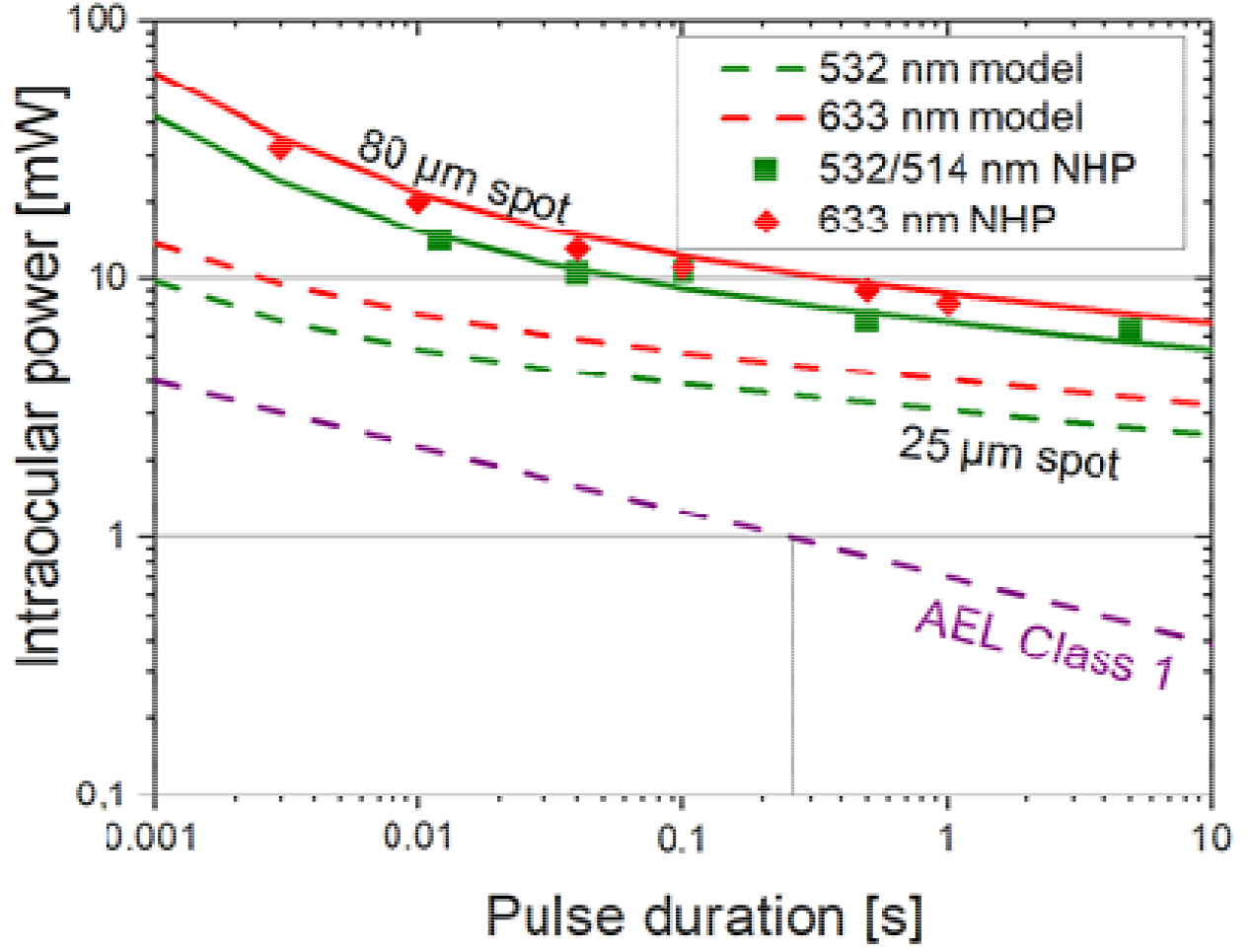
Injury thresholds obtained with NHP experiments, and a comparison with injury thresholds predicted by a computer model in the regime up to 10 seconds pulse duration (or exposure duration). Both axes are plotted in a logarithmic scale.

There is an uncertainty if the limits for the human case could be lower, as discussed by Schulmeister et al.(28) . As a conservative assumption for the human case, representing a minimum angular subtense of the retinal image of 1.5 mrad, the injury threshold was calculated for a retinal image diameter of 25 µm, shown as dashed lines. The injury thresholds were calculated for a stationary retina, which is applicable for a comparison with anesthetised experimental animals.

When applying the computer model data for a safety assessment of human exposure, it is generally accepted that the NHP thresholds, for the red wavelength range, are conservative even for highly pigmented human choroids. The AEL for Class 1 is also shown - it reaches 1 mW for a pulse duration of 0.25 s, equal to the AEL for Class 2. The injury thresholds, expressed as intraocular power, decrease with increasing pulse duration. However, this decrease occurs to a much lesser extent than the decrease in the Class 1 AEL. This results in an increasing safety margin between the injury threshold and the retinal thermal Class 1 AEL.

With the computer model it is possible to predict injury thresholds for exposure durations longer than 10 s, as shown in Figure 4 for a wavelength of 650 nm, for the spot size of 80 µm that would be consistent with NHP threshold studies, and for the conservative assumption of a 25 µm retinal spot size. The thresholds were calculated for a stationary retina, which results in damage thresholds that are much lower than would be applicable for a conscious human, where, even with intentional staring, involuntary eye movements would occur and spread the radiated area. (29). Therefore, the presence of eye movements would result in injury threshold more consistent with the regime of the 80 µm threshold curve. However, even for a theoretical 25 µm spot diameter without eye movements, the predicted injury threshold for a 200 s exposure duration is 2.5 mW, which is notably well above the exposure level of the EMMD (1 mW for a 7 mm pupil). The safety margin is further increased if the smaller pupil diameter that would normally occur during such exposures is taken into account.

**Figure 4.**
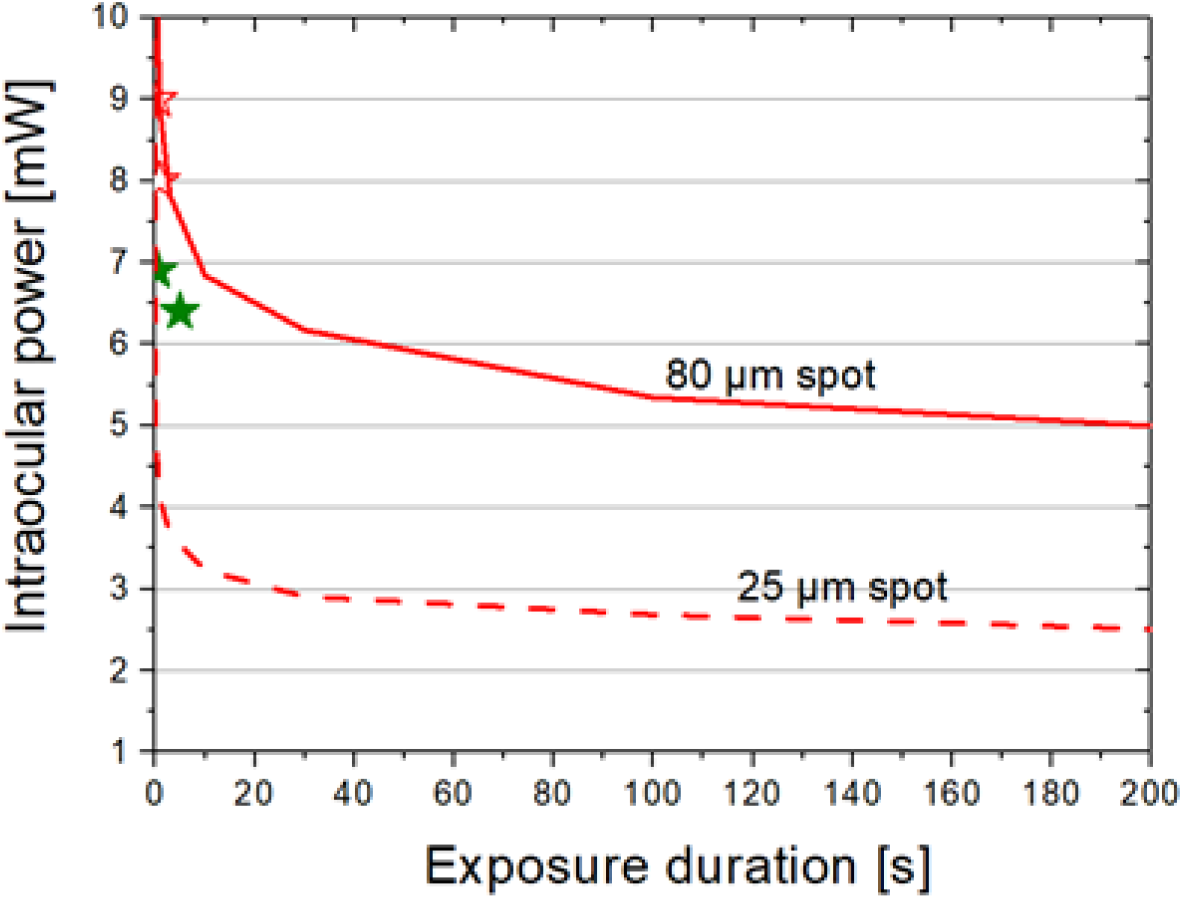
Extended range of exposure duration showing the computer model predications for a wavelength of 650 nm, neglecting eye movements. Both axes are plotted in linear scale.

### 4.4 Additivity of the photochemical hazard

Photochemical damage to cells is considered to occur as a result of photons interacting with molecular structures within the cells. In particular, there is substantial literature describing the effects of short wavelength, high energy photons, in the ultraviolet and blue end of the visible spectrum which can cause damage to the DNA helix. Earlier studies on photochemical retinal damage focused on the blue end of the spectrum following the work of Ham (15) and Lund et al. (30). This work led to the identification of a special class of photochemical retinal injury documented in the safety standards and referred to as the blue-light hazard, with a peak at 440 nm. This form of injury was described as a photic retinopathy and may explain retinal damage caused by prolonged sun-gazing rather than the more commonly used term solar burns.

More recently, there is accumulating evidence that prolonged exposure may induce photochemical changes even at longer wavelengths. Photochemical retinal injury is documented to follow the Bunsen-Roscoe law of reciprocity between irradiance and exposure duration (30, 31). This law states that if the irradiance level at the tissue is reduced by a certain factor, increasing the exposure duration by the same factor results in the same photochemical effect. The limitation of this relationship occurs at very short exposure durations and high irradiance levels, where thermal effects may dominate, and at very long exposure durations with very low irradiance levels, where biological repair mechanisms become effective. The law of reciprocity is equivalent to the experimental finding that the injury threshold, for a given wavelength, corresponds to a specific retinal radiant exposure value H_thr_ measured in J cm^-2^ that does not depend on the exposure duration t_exp_ . For a given retinal irradiance E_ret_ measured in W cm^-2^, the retinal radiant exposure H_ret_ is calculated as follows:

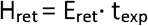

Thus, the same photochemical hazard level can be reached either when retinal irradiance is low but the exposure duration is long, or when irradiance is high but the exposure duration is short. This additivity of the photochemical hazard also applies, within limits, to exposure scenarios separated in time, such as the two daily exposures prescribed for the EMMD. The relevant quantity in this context is the total retinal radiant exposure.

For a given pupil diameter and retinal image diameter, retinal radiant exposure can be converted to the energy Q, and retinal irradiance can be converted to the power P entering the eye through the pupil. The relationship then becomes:

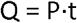

### 4.5 Multi-day exposure factor

For sufficiently long separations of the exposures, the additivity of the photochemical retinal hazard is counteracted by repair mechanisms. By determining the lesion threshold for exposures separated by 24 hours to six days, Griess & Blankenstein (32) determined a repair rate for photochemically induced retinopathy of *k* = 2.9·10^-6^ per second, equal to 0.0104 per hour. For the case where repair during the exposure phase is neglected, and assuming daily exposure, Griess and Blankenstein derived a formula that characterised the reduction factor *δ* for the threshold compared with the threshold for a single exposure. Since the maximum of treatment days for the EMMD is 5 days within a 7-day week, assuming daily exposure is a conservative assessment. In the formula, *T* is the exposure duration per day in hours, and *k* is the repair rate in hour^-1^:

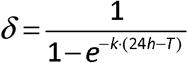

For the daily exposure duration for two EMMD treatments totalling 360 s (0.1 hour), the resulting value is *δ* = 4.5. This reduction factor is used in this discussion to account for the additive effect of exposures over multiple days.

### 4.6 Human volunteer study

In evaluating the relevance of the current classification system based on MPEs, which are derived from animals injury data, for assessing the potential hazards of the EMMD, it is useful to review an empirical study demonstrating the innate safety margins between the ED50 injury database and the corresponding MPE limits through both subjective and objective responses to exposures observed in human volunteers.

Robertson et al. in 2000 (33) reported on an experiment with human volunteers, in which laser pointers with powers up to 5 mW were used for intentional long-term retinal exposure, with participants instructed to stare directly into the beam. The laser beams had a diameter such that the total emitted power entered the eye. The beams were well collimated producing a worst-case minimum retinal image. The laser assembly was mounted on a slit lamp so that the patients’ heads were stabilised using both chin and forehead rests. Stabilizing the position of gaze was facilitated using an Amsler grid for fixation, with openings cut to allow transmission of the laser beam. Thus, the exposure scenario, and the resulting worst-case retinal image diameter, was equivalent to that produced by the EMMD when the eye accommodates to approximately 32 cm in front of the eye, generating a minimum retinal image. The laser parameters used in the experiment are summarised in Table 1.

**Table 1:** Laser parameters for the experiment with human volunteers.

|  | **Wavelength** | **Laser power** |
| --- | --- | --- |
| Laser #1 (patient #1) | 673 nm | 0.86 mW |
| Laser #2 (patient #2) | 673 nm | 1.95 mW |
| Laser #3 (patient #3) | 659 nm | 5.10 mW |

Laser pointers #1 and #2 had the lowest power level and, with a wavelength of 673 nm, have a somewhat longer wavelength than the EMMD. Therefore, the discussion here concentrates on patient #3 where the wavelength was 659 nm and the power level was measured to be 5.1 mW. This is approximately 5 times higher than the power level of the EMMD entering the eye through a 7 mm pupil. The differences in wavelength can be considered negligible. For all patients, the exposure duration was 60 seconds centrally, i.e. at the fovea. Two additional exposures lasted 5 minutes and 15 minutes, and were incident 5° above and below the fovea, respectively.

In the paper the volunteers reported that for the 60 s exposure at 5.1 mW, they experienced no subjective visual effects other than a pink after-image, which lasted approximately five minutes before fading. Their visual acuity returned to 20/20, within the 3 to 4 minutes after exposure. When examined at 4 days and 15 days after exposure, there was no evidence of phtopsias, visual field defects, or fundus abnormalities that could be attributed to the effects of viewing the laser pointer. These observations are particularly important because, in most reported cases of low-level laser injury, patients present with self-reported visual disturbances. In the discussion section of their paper, summarizing the findings for all three patients, the authors stated that they observed neither acute laser injuries of the retina nor any clinical evidence of delayed photic retinopathy. Other than the reported transient after images, they were unable to document any functional, ophthalmoscopic, fluorescein angiographic, or histologic evidence of damage to the human retina after exposure to 1, 2, and 5 mW for durations of up to 15 minutes.

The absence of detectable retinal injuries indicates that these findings apply to both thermally and photochemically induced retinopathy.

This study is consistent with the thermal threshold data shown in Figure 4, supporting that the computer model data is conservative. Thermally induced cell damage is non-linear with temperature(34) . A power level of 1 mW, produces approximately one fifth of the temperature rise resulting from a power level of 5 mW. The thermal hazard of a power level of 1 mW can therefore be considered sufficiently low that thermally induced retinal injury would not be expected. In addition, experience from skin exposure studies indicates that there is negligible additivity of thermal hazard for daily exposures. The margin between the 5 mW power levels used in the Robertson study and 1 mW exposure from the EMMD (for a 7 mm pupil) also accommodates potential variability in choroidal pigmentation.

For comparison of the EMMD exposure with the Robertson study in relation to photochemical hazards, the law of reciprocity can be applied to power levels measured in mW as follows. In the Robertson study, 5 mW of radiant power for an exposure duration of 15 minutes did not result in detectable retinopathy. This corresponds to a potential photochemical hazard equivalent to 1 mW of radiant power for an exposure duration of 75 minutes (5 mW/1 mW × 15 minutes). In terms of energy, 5 mW × 15 min corresponds to the same radiant exposure as 1 mW × 75 min.

Since the daily exposure duration for the EMMD is 6 minutes, the margin between the Robertson exposure that produced no detectable injury and the EMMD for a 7 mm pupil diameter is 75 min/6 min = 12.5. This ratio increases further when more realistic pupil diameters are considered. For example, a 4 mm pupil diameter results in a ratio of approximately 38.

Exposures in the Robertson study took place within a single day, and no further exposures were applied. Daily exposure for an unlimited period of time can be accounted for by applying the reduction factor *δ* = 4.5 derived earlier. For a 7 mm pupil diameter, the ratio of 12.5 divided by 4.5 leaves a ratio of 2.8 relative to exposure levels that did not produce any detectable effect, i.e. the margin to result in a relevant effect is higher, but unknown. For the more realistic, although still conservative, 4 mm pupil diameter, the ratio of 38 reduced by *δ* = 4.5 results in a factor of 8.5. It may therefore be argued that this margin also accounts for potential population variability in retinal sensitivity and injury thresholds, including potential differences between paediatric and adult eyes, although such differences have not been observed to date.

### 4.7 Strengths and Limitations

This study has several strengths, including direct measurement of device-specific emission parameters in an accredited laboratory, integration of established thermal and photochemical injury models, and comparison with empirical human exposure data to provide a comprehensive safety assessment. The analysis was intentionally based on conservative, worst-case assumptions, including maximal pupil diameter, minimal retinal image size, and absence of eye movements, thereby strengthening the robustness of the safety margins. However, several limitations should be acknowledged. The study did not include prospective clinical safety data in treated paediatric populations, and conclusions rely in part on extrapolation from experimental models and adult volunteer studies. In addition, inter-individual variability, including differences in ocular pigmentation and potential photosensitivity, may influence susceptibility to retinal effects. These considerations highlight the value of continued clinical monitoring and future studies in real-world populations to further strengthen the evidence base.

## CONCLUSIONS

Given that there are no specific safety standards designed to address the exposure parameters of repeated low-level red-light therapy, intended to retard or stop progressive myopia in children, the present study compared the measured exposure parameters of one such system (the EMMD) with the limits defined in current codes of practice. Based on measurements obtained from EMMD and careful consideration of these standards, the device falls within the Class 3R category, but it should be noted that it only marginally exceeds the Class 2 limit. It should also be emphasised that these codes of practice were not designed for therapeutic applications, where consideration of risk versus clinical benefit is of greater importance.

Comparison with data from a human volunteer study, in which participants were intentionally exposed to laser pointers with similar but higher exposure levels, revealed no subjective or objective evidence of retinal injury. This finding illustrates the substantial safety margins incorporated into the standards, ensuring that experimentally determined injury thresholds (ED50 values) remain well separated from the maximum permissible exposure (MPEs) limits defined in the codes of practice.

On the basis of these considerations, we conclude that the EMMD can be considered safe when used according to the exposure regimes defined by the manufacturer’s software controls. However, it cannot be excluded that certain individuals may have increased photosensitivity, for example, due to the use of photosensitizing medication, such as those used in photodynamic therapy. As is common in radiation safety, such individuals should take appropriate precautions or avoid such treatments.

## Data Availability

All data produced in the present work are contained in the manuscript

## AUTHOR CONTRIBUTIONS

Dr. Schulmeister had full access to all of the data in the study and takes responsibility for the integrity of the data and the accuracy of the data analysis. Dr. Schulmeister and Prof. Marshall were equally involved in concept and design of the study and preparing and critically reviewing the manuscript.

